# The closer to the Europe Union headquarters, the higher risk of COVID-19? Cautions regarding ecological studies of COVID-19

**DOI:** 10.1101/2020.04.23.20077008

**Authors:** Shuai Li, Xinyang Hua

## Abstract

Several ecological studies of the coronavirus disease 2019 (COVID-19) have reported correlations between group-level aggregated exposures and COVID-19 outcomes. While some studies might be helpful in generating new hypotheses related to COVID-19, results of such type of studies should be interpreted with cautions. To illustrate how ecological studies and results could be biased, we conducted an ecological study of COVID-19 outcomes and the distance to Brussels using European country-level data. We found that, the distance was negatively correlated with COVID-19 outcomes; every 100 km away from Brussels was associated with approximately 6% to 17% reductions (all P<0.01) in COVID-19 cases and deaths in Europe. Without cautions, such results could be interpreted as the closer to the Europe Union headquarters, the higher risk of COVID-19 in Europe. However, these results are more likely to reflect the differences in the timing of and the responding to the outbreak, *etc*. between European countries, rather than the ‘effect’ of the distance to Brussels itself. Associations observed at the group level have limitations to reflect individual-level associations – the so-called ecological fallacy. Given the public concern over COVID-19, ecological studies should be conducted and interpreted with great cautions, in case the results would be mistakenly understood.

## Introduction

The year of 2020 has been hallmarked by the coronavirus disease 2019 (COVID-19). As of 22 April 2020, the COVID-19 pandemic had been reported to cause more than 2.4 million cases and 169,006 deaths worldwide^1^.

Many studies have been conducted to understand this ‘unknown’ new disease. Some studies found that regional aggregated exposures (i.e., Bacillus Calmette-Guerin vaccination, air quality index) were correlated with regional aggregated COVID-19 epidemic measures (i.e., numbers of cases and deaths)^2–6^, with implications that high levels of certain vaccinations and lower levels of certain air pollutants would protect from COVID-19. These studies typically investigate if there is a certain pattern in the heatmap (like Figure 1) of COVID-19 epidemic, and if the pattern (if there is one) is correlated with the patterns in the heatmaps of potential exposures. Such type of studies using group-level data, rather than individual-level data, is called ecological studies^7^. While some ecological studies might be helpful in generating new hypotheses related to COVID-19, their results should be interpreted with cautions, as ecological studies by nature are sometimes more vulnerable to bias than studies using individual-level data; see below.

**Figure 1.**
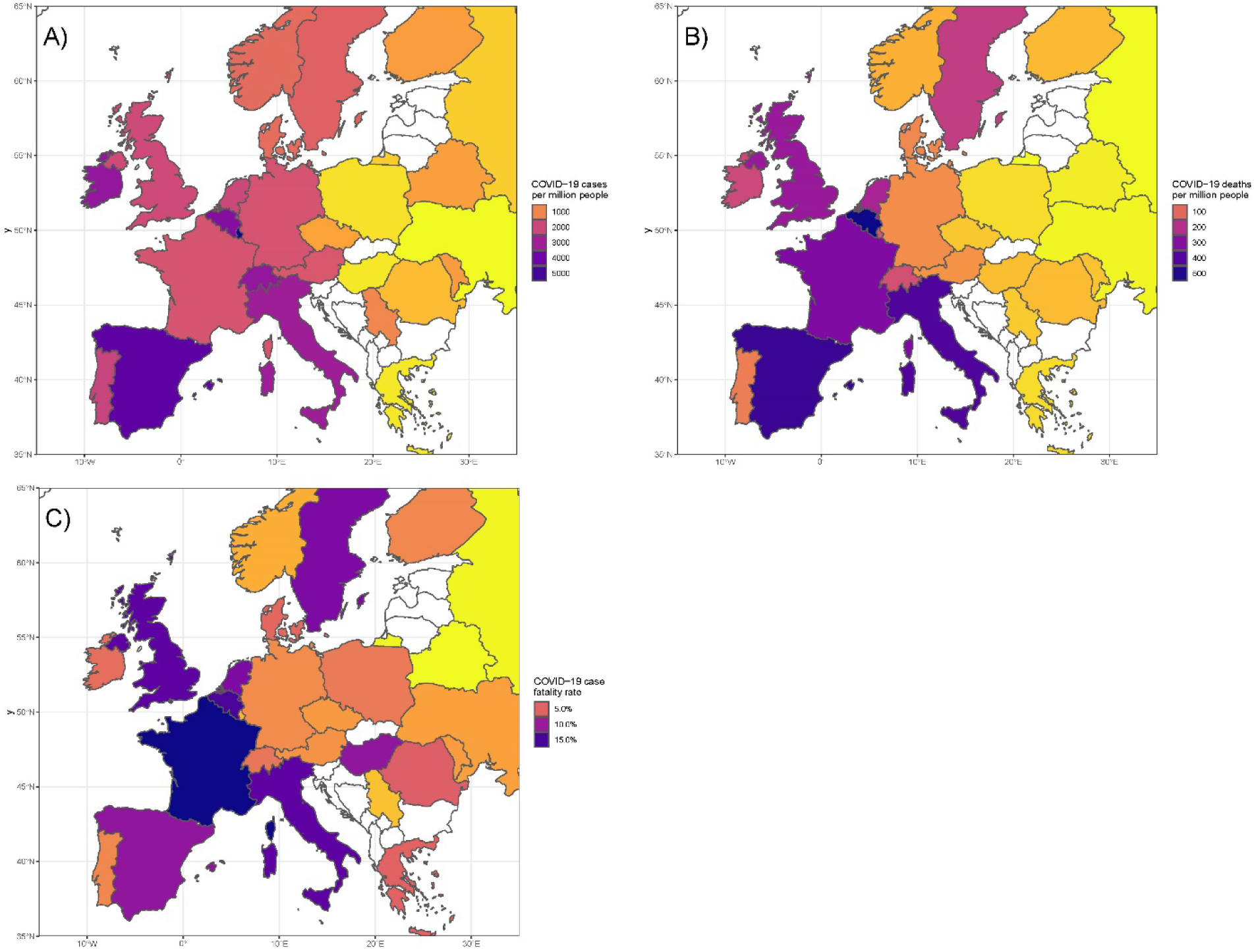
The geographic distributions of the COVID-19 epidemic measures for European countries. Countries coloured blank were excluded from analysis due to having fewer than 2000 cumulative cases. A) The number of COVID-19 cases per million people; B) The number of COVID-19 deaths per million people; C) The COVID-19 case fatality rate

To illustrate how ecological studies and relevant results could be biased, we presented an ecological study of investigating the relationships between COVID-19 epidemic measures and the distance to Brussels, where the Europe Union (EU) headquarters located, in Europe.

## Methods

We obtained the COVID-19 data for European countries from the European Centre for Disease Prevention and Control (ECDC) (https://www.ecdc.europa.eu/en/publications-data/download-todays-data-geographic-distribution-covid-19-cases-worldwide). The data file provided by the ECDC contains country-specific daily numbers of COVID-19 cases and deaths, and population size in 2018. To minimise the results being impacted by small numbers of cases, we included countries with at least 2,000 cumulative cases (up to and including 22 April 2020) only. A total of 26 countries were included (Figure 2). For each country, we calculated three COVID-19 epidemic measures: number of cases per million people calculated using the number of cumulative cases and population size, number of deaths per million people calculated as the number of cumulative deaths and population size, and case fatality rate defined as the proportion of cumulative deaths in cumulative cases. Figure 1 shows that, Italy, Spain, and North-western and Western European countries had more serious epidemics of COVID-19 than the other European countries. There is a pattern consistently showed by these maps – European countries with serious epidemics appear to be around Belgium (and Belgium is one of the countries with the highest values in the three COVID-19 epidemic measures). Inspired by this pattern, we further investigate if the COVID-19 epidemic measures correlate with the distance to Belgium for these countries. For each country, the distance was defined as the direct distance (as the crow flies) from its capital to Brussels, and measured using an online tool (https://www.freemaptools.com/how-far-is-it-between.htm).

**Figure 2.**
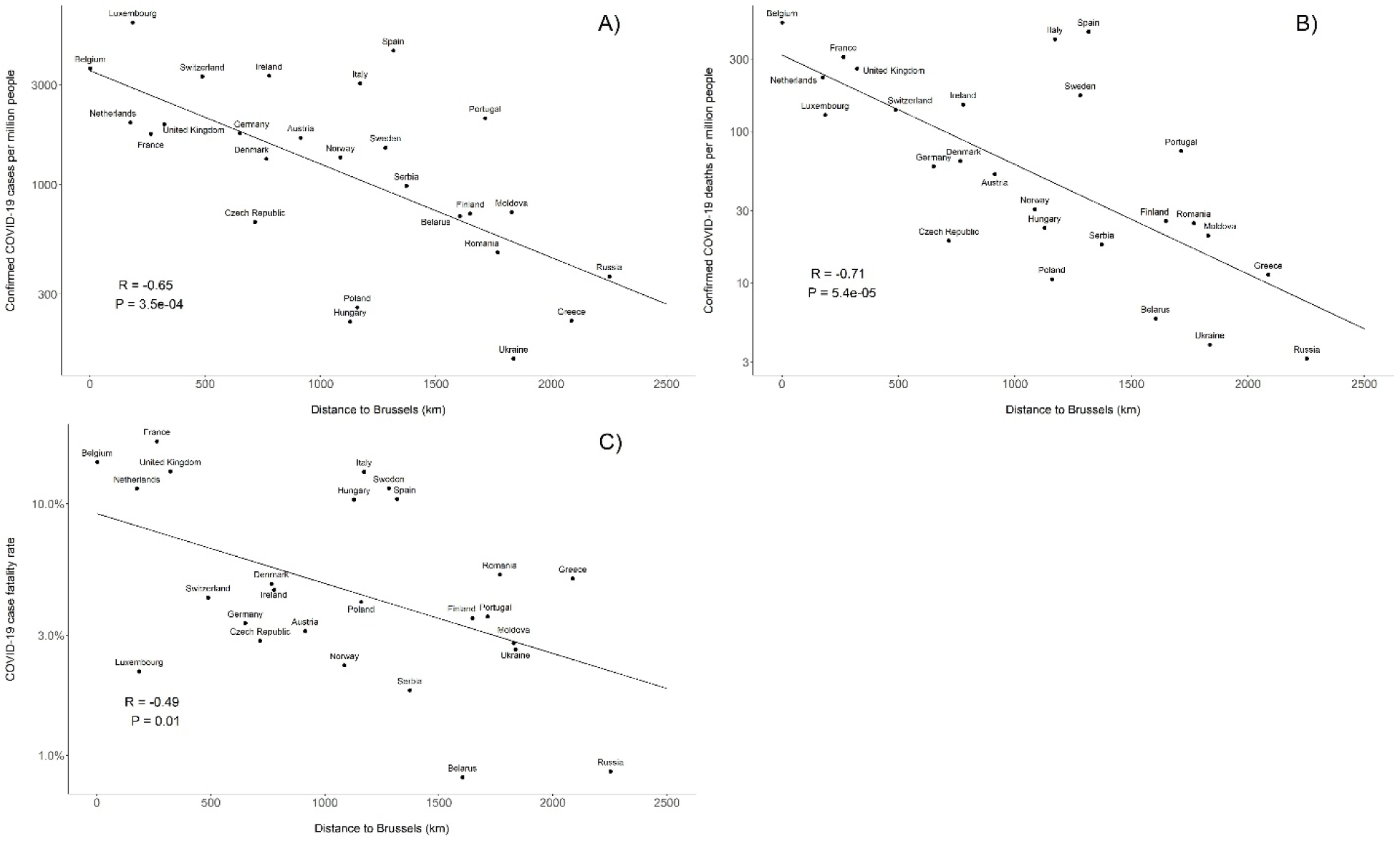
Correlations between COVID-19 epidemic measures and the distance to Brussels for European countries. The y-axis is in log scale. R is the Pearson correlation coefficient and P is the corresponding P-value. A) Correlation for the number of COVID-19 cases per million people; B) Correlation for the number of COVID-19 deaths per million people; C) Correlation for the COVID-19 case fatality rate

We investigated the correlations between the distance and each of the three outcomes (log-transformed) using Pearson correlation coefficient. We estimated the ‘effect’ of the distance on COVID-19 outcomes using a log-linear regression model.

## Results

All the three COVID-19 outcomes were negatively correlated with the distance to Brussels (Figure 1); the correlation coefficient ranged from −0.5 to −0.7, and all the P-values were less than 0.01.

From the log-linear regression, every 100km away from Brussels was associated with 10.3% (95% confidence interval [CI], 5.4% to 15.1%) reduction in confirmed COVID-19 cases per million people, with 16.7% (95% CI, 10.0% to 23.3%) reduction in confirmed COVID-19 deaths per million people, and 6.4% (95% CI, 1.8% to 11.0%) reduction in COVID-19 case fatality rate.

## Discussion

From our example of an ecological study, we found that for European countries their distance to Brussels was negatively correlated with their COVID-19 epidemic measures, and all the correlations were statistically significant. If no cautions were given to our study, such results could be interpreted as the closer to the EU headquarters, the higher risk of COVID-19 in Europe, with an implication that Europeans might need to move away from Brussels to get protected from COVID-19 – a conclusion and implication with limited plausibility.

Ecological bias, or ecological fallacy, is the major limitation of ecological studies in making causal inference. This bias is usually interpreted as the failure of the observed association at the group level to reflect the biological effect at the individual level^7^. While a strong individual-level effect of an exposure on an outcome could result an effect at the group level, i.e., a population with more smokers tend to have more lung cancers compared with a (comparable) population with fewer smokers, the reverse is not always held. One source for ecological bias is that the group-level measures do not necessarily reflect the measures at the individual level, as the latter are not measured at all. Although in our example, the country distance could reflect to some extent the individual-level distance to Brussels of the people living in that country, the COVID-19 epidemic measures were not at the individual level, and there are other factors could bias the association; see below.

It is argued that the purpose of some ecological studies is to make inference for the ecological effects of an exposure on group rates, rather than to make inference for the biological effects on individual disease risks^8^. However, such two purposes tend not to be explicitly differentiated by some studies, and results from studies with ecological purposes are vulnerable to be mistakenly interpreted as biological effects. Furthermore, even with the ecological purpose, cautions are still needed. Ecological bias could not only arise from the bias at the individual level, but could also arise from confounders between groups and effect modification by group^9^. The latter two are unique to ecological studies; therefore, ecological studies are sometimes more vulnerable to bias than individual-level studies. Observed ecological associations could be due to these biases. In terms of our example, there are considerable differences in the epidemic stage, responding to the outbreak, test capacity, healthcare system, population structure, *etc*. between European countries that could confound the association at the country level. Our observed ecological association is more likely to reflect the impacts of these differences on the varied COVID-19 epidemic measures between the studied European countries, than the ‘effect’ of the distance to Brussels itself.

Nevertheless, ecological studies indeed have some values. Professor Neil Pearce^10^ thinks there are two reasons for the modern revival of ecological studies: 1) population-level studies is valuable in identifying important public health problems and in generating hypotheses about the potential causes of these problems (as he pointed out, some discoveries about the causes of cancer could be attributed to the hypotheses generated by internationally comparing cancer incidences conducted half a century ago); and 2) the recognition that some risk factors may also impact individual disease risk at the population level. It might be too immature to conclude at this stage whether the emerging ecological studies could contribute to our understanding of COVID-19 from the two perspectives above. The current studies might inspire some further studies to research in depth to provide more evidence for supporting, or falsifying, relevant hypotheses.

Understanding the differences in health conditions between populations is the driving force of the development of epidemiology; however, modern epidemiology tends to more focus on the individual level. Comparing group-level data could bring the public-health orientation back to epidemiology, but simply conducting ecological studies would have little help^7^. Studies systematically comparing country-level COVID-19 outcomes with appropriate designs could potentially provide insights into the possible factors contributing to the varied epidemic across countries, and provide policy evidence for responding similar epidemics in the future.

Given the public concern over COVID-19, any ‘novel/striking finding’ from ecological studies would have the potential of becoming eye-catching headlines and attract considerable media coverage, and be easily interpreted problematically. Findings reported without peer-review potentially have more profound influences. Health policy and clinical practice might be minimally influenced by these findings; however, considerable influences on the public without relevant expertise are not unlikely. Therefore, we suggest that health researchers should take cautions when conducting ecological studies of COVID-19, and sharing and discussing relevant results.

## Data Availability

The data were accessed from public sources.

## Author’s contributions

SL conceived the study and analysed the data. SL and XH wrote the manuscript.

## References

1. WHO. Coronavirus disease 2019 (COVID-19) Situation Report – 93 [Available from: https://www.who.int/docs/default-source/coronaviruse/situation-reports/20200422-sitrep-93-covid-19.pdf?sfvrsn=35cf80d7_4 accessed 23 Apr 2020.

2. Miller A, Reandelar MJ, Fasciglione K, et al. Correlation between universal BCG vaccination policy and reduced morbidity and mortality for COVID-19: an epidemiological study. medRxiv 2020:2020.03.24.20042937. doi: 10.1101/2020.03.24.20042937

3. Berg MK, Yu Q, Salvador CE, et al. Mandated Bacillus Calmette-Guérin (BCG) vaccination predicts flattened curves for the spread of COVID-19. medRxiv 2020:2020.04.05.20054163. doi: 10.1101/2020.04.05.20054163

4. Wu X, Nethery RC, Sabath BM, et al. Exposure to air pollution and COVID-19 mortality in the United States. medRxiv 2020:2020.04.05.20054502. doi: 10.1101/2020.04.05.20054502

5. Travaglio M, Popovic R, Yu Y, et al. Links between air pollution and COVID-19 in England. medRxiv 2020:2020.04.16.20067405. doi: 10.1101/2020.04.16.20067405

6. Setti L, Passarini F, De Gennaro G, et al. The Potential role of Particulate Matter in the Spreading of COVID-19 in Northern Italy: First Evidence-based Research Hypotheses. medRxiv 2020:2020.04.11.20061713. doi: 10.1101/2020.04.11.20061713

7. Rothman KJ, Greenland S, Lash TL. Modern epidemiology. 3rd. Philadelphia: Lippincott Williams & Wilkins 2008: 26–30.

8. Morgenstern H. Uses of ecologic analysis in epidemiologic research. American journal of public health 1982;72(12):1336–44. doi: 10.2105/ajph.72.12.1336 [published Online First: 1982/12/01]

9. Greenland S, Morgenstern H. Ecological bias, confounding, and effect modification. International journal of epidemiology 1989;18(1):269–74. doi: 10.1093/ije/18.1.269 [published Online First: 1989/03/01]

10. Pearce N. The ecological fallacy strikes back. Journal of Epidemiology and Community Health 2000;54(5):326–27. doi: 10.1136/jech.54.5.326

